# RNA expression classifiers from a model of breast epithelial cell organization to predict pathological complete response in triple negative breast cancer

**DOI:** 10.1101/2021.02.10.21251517

**Authors:** Joan W. Chen, Ryan P. Russell, Trushna Desai, Mary Fiel-Gan, Varun Bhat, Maria de Fátima Dias Gaui, Luis Claudio Amendola, Zilton Vasconcelos, Adam M. Brufsky, Marcia V. Fournier, Susan H. Tannenbaum

## Abstract

Pathological complete response (pCR) to neoadjuvant chemotherapy (NAC) is correlated with better outcomes for breast cancer, especially for triple negative breast cancer (TNBC). We developed RNA expression classifiers from a model of breast epithelial cell organization to predict which patients will achieve pCR to NAC, and which will have residual disease (RD). An exclusive collection of retrospective formalin-fixed, paraffin-embedded (FFPE) pretreatment biopsies from 222 multi-institutional breast cancer patients treated with NAC, including 90 TNBC patients, were processed using standard procedures. A novel strategy using machine learning algorithms and statistical cross-validation were used to develop predictive classifiers based on AmpliSeq differential gene expression analysis of patient samples. Two RNA expression classifiers of 18 genes and 15 genes applied sequentially to the total cohort, classified patients into three distinct classes which accurately identified 83.75% of pCR and 86.62% of RD patients in the total population, and 92.10% of pCR and 80.77% of RD patients in the TNBC subset. This new approach identified a subset of TNBC patients predicted to have RD showing significantly higher levels of Ki-67 expression and having significantly poorer survival rates than the other TNBC patients. Stratification of patients may allow identification of TNBC patients with the worst prognosis prior to NAC, allowing for personalized treatments with the potential to improve patient outcomes.

**Statement of Significance:** Stratification of TNBC patients by prognosis prior to NAC, may allow for more personalized treatment approaches with the potential to improve patient outcomes and reduce toxicity.

## Introduction

Breast cancer incidence has not changed significantly over the last few decades. Mortality trends have significantly improved with 79% survival from 1984 through 1986 and 91% survival in 2008-2014 in accord to the American Cancer Society annual report. This improvement in survival is primarily seen in hormone receptor positive and HER-2 positive subtypes. The triple negative subtype has lagged behind the others and until recently was limited to non-targeted treatments [1].

A seminal meta-analysis of multiple trials published in 2014 confirmed that patients who have achieved a pathological complete response (pCR) to chemotherapy given prior to surgery, neoadjuvant chemotherapy (NAC), have improved survival [2]. They demonstrated that the more aggressive subtypes, triple negative and HER-2/neu positive, had increased frequencies of pathological complete response with improvement in event free and overall survival. Outside of a clinical trial setting, this was confirmed in a real-world setting showing that pCR can be applied as a surrogate endpoint for survival especially in triple negative breast cancer (TNBC) [3].

Unfortunately, TNBC which represents about 15-20% of all breast cancer, is a heterogeneous disease with distinct molecular subtypes. In 2013 an initial publication looked at the differential response retrospectively to NAC in 130 patients stratified by the 7 molecular subtypes defined for TNBC at that time. The basal-like 1 subtype had the highest pCR of 52% and the lowest was seen in the basal-like 2 at 0% [4]. A revised classification defined 4 TNBC subtypes [5] with significant differences in presentation, response to treatment, sites of metastatic disease and prognosis [6]. There have been innumerable publications looking into revised molecular classifications of TNBC and response to NAC therapy which is summarized by Garrido-Castro in 2019 [7].

In unselected patients with TNBC, BRCA mutations were found in approximately 20% of patients. Due to deficiencies in BRCA-associated DNA repair mechanisms, DNA damaging agents play a role in TNBC. This is demonstrated in studies utilizing platinum-based regimens as well as poly (ADP-ribose) polymerase PARP inhibitors in both sporadic patients with TNBC and those with germline mutations in BRCA 1 and 2 [[8], [9]]. Additional studies also implicate the tumor microenvironment, in particular tumor infiltrating lymphocytes and PD-L1 status, in outcomes and responses to chemotherapy [[10], [11]]. Multiple studies in patients with TNBC in the metastatic and neoadjuvant setting have demonstrated impressive increases in pCR [[12], [13]], The development of new strategies for the treatment of TNBC holds promise for patients and expression biomarkers can help guide which patients may benefit the most of these new approaches with a goal of escalating or deescalating treatment as appropriate.

The unresolved question remains, is there a biomarker or biomarker panel that can assist in determining response to standard and still currently recommended (NCCN Guidelines May 2020; NCCN.org) anthracycline/taxane based chemotherapy? Can we do better with our choices in the neoadjuvant setting, with greater certainty in response and reduction in utilization of increased numbers of drugs which result in both financial and personal toxicity?

We have previously described a novel strategy to develop RNA expression classifiers using published Affymetrix microarray breast cancer datasets to predict which patients are likely to achieve pCR or residual disease (RD) to standard NAC [14]. The analyses focused on 325 biomarkers derived from an experimental model of non-malignant breast epithelial cell organization in three-dimensional laminin-rich extracellular matrix that correlated with breast cancer clinical outcomes [[15]-[17]], 23 TNBC-related genes [18], and a unique strategy using a sequential application of machine learning algorithms and statistical analyses to select and rank informative genes. The results showed that in addition to stratifying patients into pCR and RD classes, the rational for sequential application of RNA expression classifiers identified a subset of RD patients with the worst survival rates.

In the current study, we validated the same methodology on a new validation cohort of retrospective breast cancer samples collected from four different medical centers and hospitals. Using next generation sequencing, which can be utilized in the clinical setting, the results showed that the sequential application of 2 RNA expression classifiers stratified responders from non-responders to standard of care NAC with high sensitivity and specificity. These classifiers could maximize utilization of the standard regimen in those that respond, reserving more novel and intensive therapy for those who are predicted non-responders. Additionally, this study confirmed the existence of extremely poor prognosis class of TNBC patients with RD, who have the worst relapse free survival, highest initial stage, express high levels of the proliferation marker Ki67 mRNA, and lower androgen receptor (AR) mRNA. More intensive and experimental therapies could be reserved for this predefined TNBC patients with worse prognosis.

## Results

### Description of our patient population

Two hundred and eighty-seven breast cancer patient samples were obtained from multiple clinical sites including the University of Connecticut Health Center (UConn Health), Hartford Hospital, Fernandes Figueira Institute/FIOCRUZ (Brazil) and the MT Group. All samples were obtained under IRB-approval and waived from consent. Breast tumor biopsy and lymph node biopsy (when available) were collected prior to patients receiving neoadjuvant anthracycline-taxane-based chemotherapy and RNA was extracted from the resulting Formalin-Fixed Paraffin-Embedded (FFPE) biopsy sections. A total of 222 clinical samples which passed all quality control checkpoints were included in the subsequent analysis (**Table 1)**. The 222 samples cohort are composed of stage I-IV newly diagnosed invasive breast cancer treated with standard NAC incorporating a taxane, an anthracycline, and cyclophosphamide (AC-T). Based on the IHC reports the cohort included 31.1% of patients with ER-positive/HER2-negative tumors and 40.5% with TNBC. **Table 1** shows patient demographics and histopathological information. 73.3% of TNBC patients were grade 3, with 45% of grade 3 patients achieving a pCR.

**Table 1:**
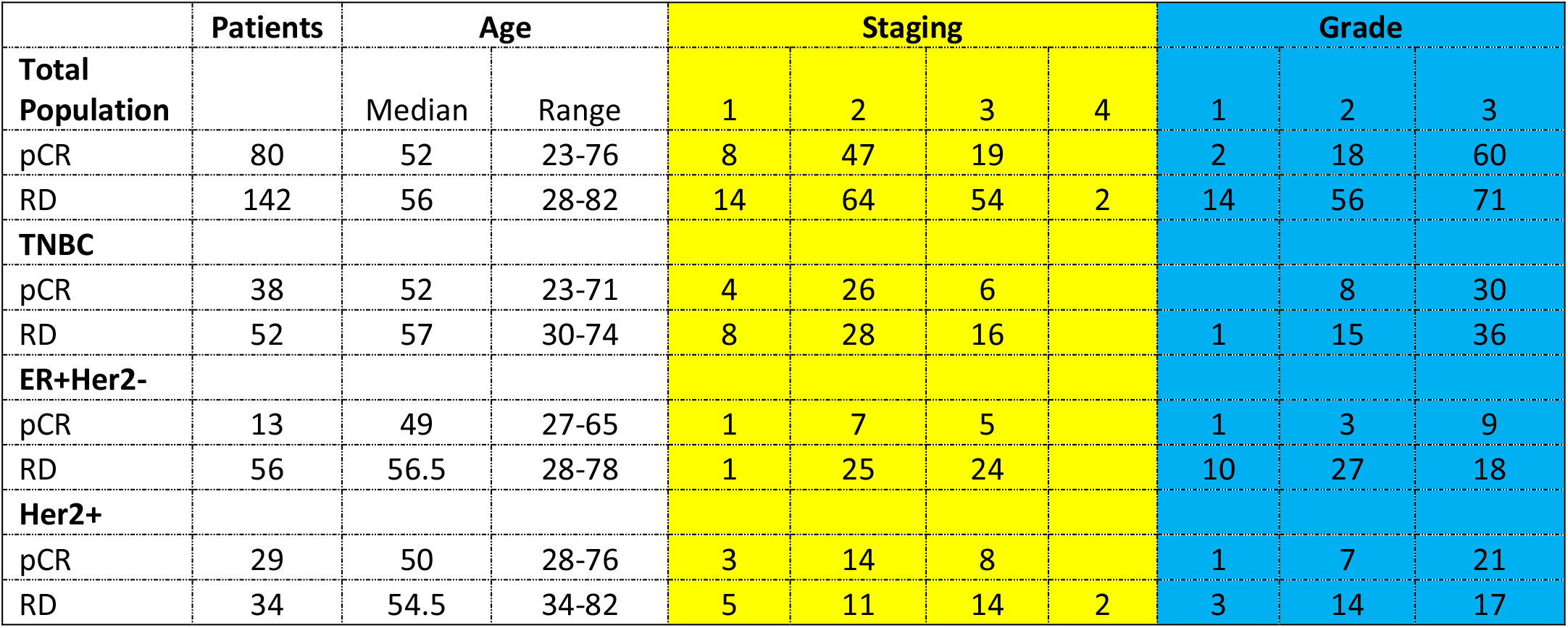
Demographics of patients and tumor characteristics in the study

Cluster analysis of all 222 samples with a previously described panel of 348 biomarkers, which includes 325 novel RNA markers [16] and 23 TNBC markers [18] revealed while clustering gives a visual graphics of patient distribution trends based on gene expression patterns, it did not provide enough resolution to provide significant class predictions (Figure S1).

### Generation of the Two pCR Prediction Classifiers

We used a previously described methodology to develop and apply RNA expression classifiers sequentially to the patient cohort to stratify patients based on clinical outcomes [14]. **Figure 1** shows diagram of model development and sequential application of classifiers to the patient population.

**Figure 1:**
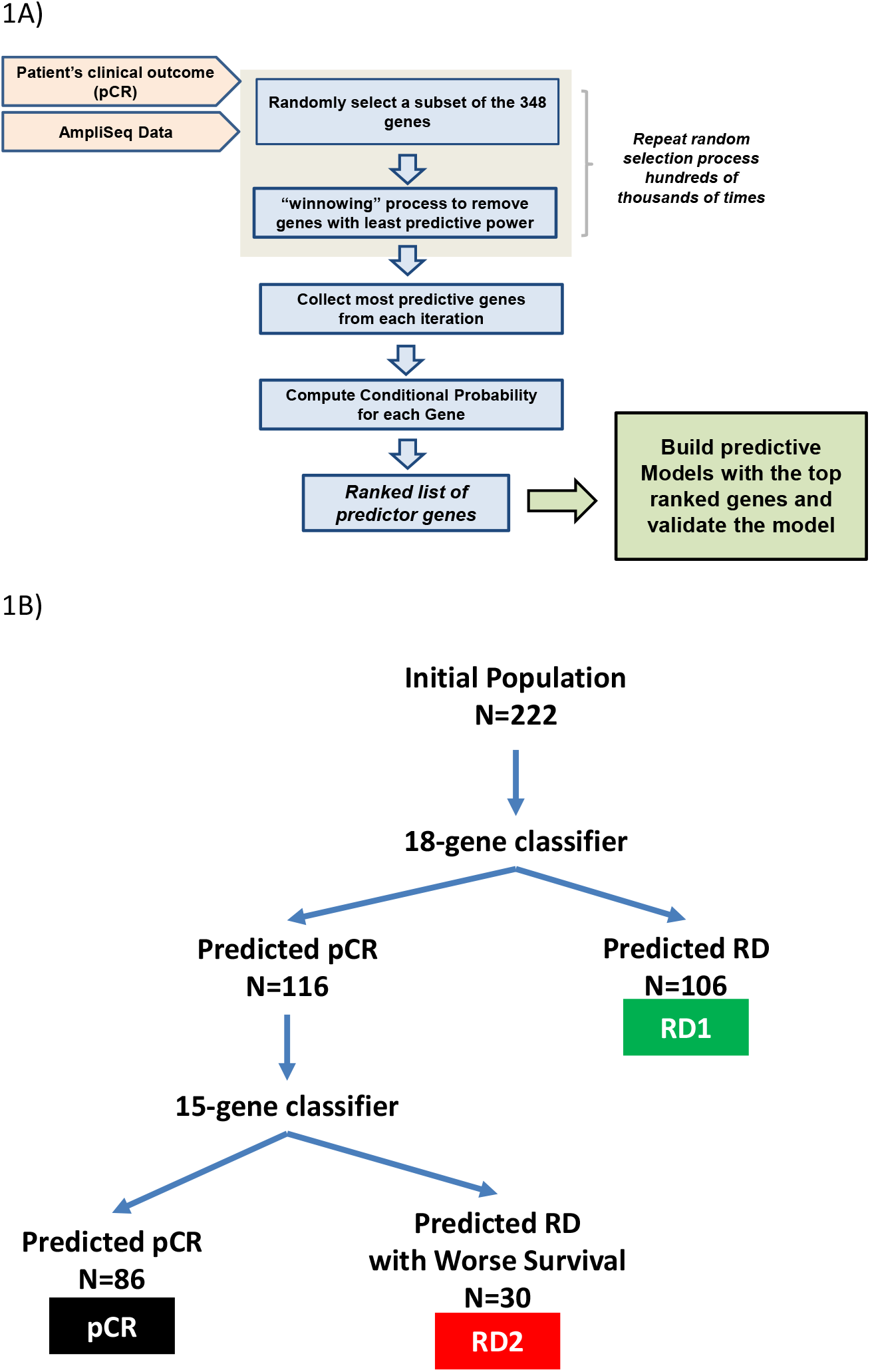
A) diagram showing the flow of the machine-learning algorithms to build pCR-predictive models using a two-step process: 1) Select the best genes for the model and 2) successively eliminate genes that are least useful to the classifier performance. B) Diagram showing sequential application of 18-gene and 15-gene classifiers to the dataset.

We combined bias-reduction generalized linear model (brglm) and bootstrapping of samples and genes to identify genes that have the most predictive values. It was found that the occurrences of genes after large number of iterations varied quite significantly (Figure S2). Genes occurred at higher frequencies after 10,000 iterations of selection are assumed to have better prediction of the clinical outcome than the genes with lower frequencies.

Five-fold cross-validation analysis with various number of top-ranked genes and bootstrapped training samples showed that the Area Under the Curve (AUC) values, a metric used to evaluate model performance, kept improving when more genes were incorporated into the prediction models (Figure 2A). However, it was not seen when the bootstrapped testing samples were used. The earliest AUC peak (first plateau) was identified around the 18 genes (Figure 2B), suggesting that the combination of the 18 genes yield as good as it gets pCR predictive models.

**Figure 2:**
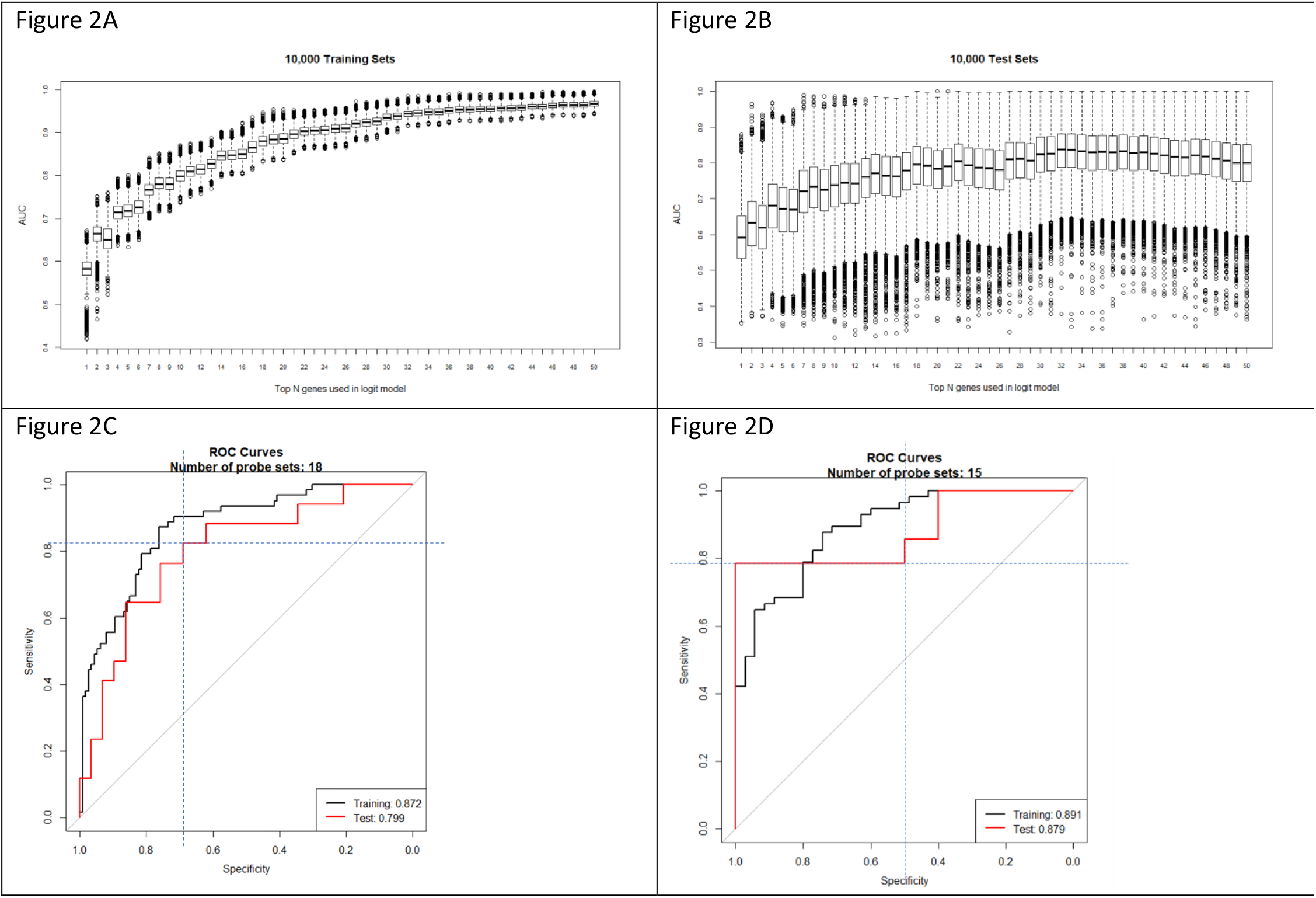
Outputs of the machine-learning algorithm run. A) Boxplot of the AUC values with the given numbers of top-ranked genes in the models from five-fold cross-validation for 10,000 time using the 80% of the training data. B) Boxplot of the AUC values with the given numbers of top-ranked genes in the models from five-fold cross-validation for 10,000 time using the 20% of the testing data. C) ROC showing the sensitivity and specificity of the predictive model with the 1^st^ 18-gene classifier. D) C) ROC showing the sensitivity and specificity of the predictive model with the 2^nd^ 15-gene classifier.

High performing models were further validated using ROC curve with the 46 testing samples which were set aside and were not used during the model development (Figure 2C). Once again, after comparing among several ROC curves with similar overall AUC values, the 18-gene model appears to yield the best combination of sensitivity and specificity.

To reduce the numbers of false positives detected among the 222 patients, we repeated the modeling process with the genes from the 348 biomarkers, but excluding the 18 genes in the first classifier, and only using the samples that were predicted as pCR by the first classifier, to develop a second 15-gene classifier (Figure 2D). The use of two sequential classifiers containing different genes allowed us to combine a general predictor for all disease subtypes with a second predictor that improves the classification of TNB patients.

Genes from the two classifiers (Table S1) are involved in various biological processes including transcriptional regulation, cell signaling, and mRNA and protein transport. For example, genes including CAPRIN2, DVL1, EHF, LRP8, and NOC2L are involved in the transcriptional regulation of gene expression; and genes including BCAR3, DVL1, LRP8, PGK1, STK17A and TTK are involved in protein phosphorylation and cell signaling pathways. DVL1 gene encodes a cytoplasmic phosphoprotein that regulates cell proliferation, and has elevated expression in glioma, hepatocellular carcinoma, as well as in sensitizing paclitaxel-resistant human ovarian cancer cells via AKT/GSK-3β/β-catenin signaling [[19], [20]]. BCAR3 is a protein that is associated with a number of tumors. It encodes a protein with a putative Src homology 2 (SH2) domain and participates in the EGF mitogenic signaling pathway which leads to estrogen-independent cell cycle progression and cell proliferation of breast cancer cells [21]. In addition, it has been shown that BCAR3 can be used as a potential biomarker, and is an independent prognostic factor for Multiple myeloma [22]. Genes involved in the extracellular matrix, such as MFAP4, were also identified in the classifiers. The extracellular matrix (ECM) protein microfibrillar-associated protein 4 (MFAP4) is has been shown to bind to ECM fibers including collagen, tropoelastin, and fibrillin *in vitro*, and is involved in cell adhesion or intercellular interactions and in disease-related tissue remodeling [23]. MFAP4 has been shown recently as a novel biomarker in human cancers [24], as well as in hepatic fibrosis to identify high-risk patients with severe fibrosis stages in hepatitis C [25]. These findings were consistent with the original selection of the 325 biomarkers based on an experimental model of breast epithelial cells [16].

### Model Performance and Description of a Subset of TNBC with Worse Prognosis

The predictive scores obtained from the 18-gene and 15-gene models were plot on the 2-dimentional scatter plots for the total 222-patient population, as well as for the 90-patient TNBC population. As shown in Figure 3, the predicted pCR samples are concentrated in the upper right quadrant. On the other hand, patients predicted RD are distributed over the other three quadrants. The population of RD patients was further stratified by the second 15-gene classifier resulting in 2 predicted RD populations (RD1 and RD2) representing a distinct biology as defined by the 2 classifiers.

**Figure 3:**
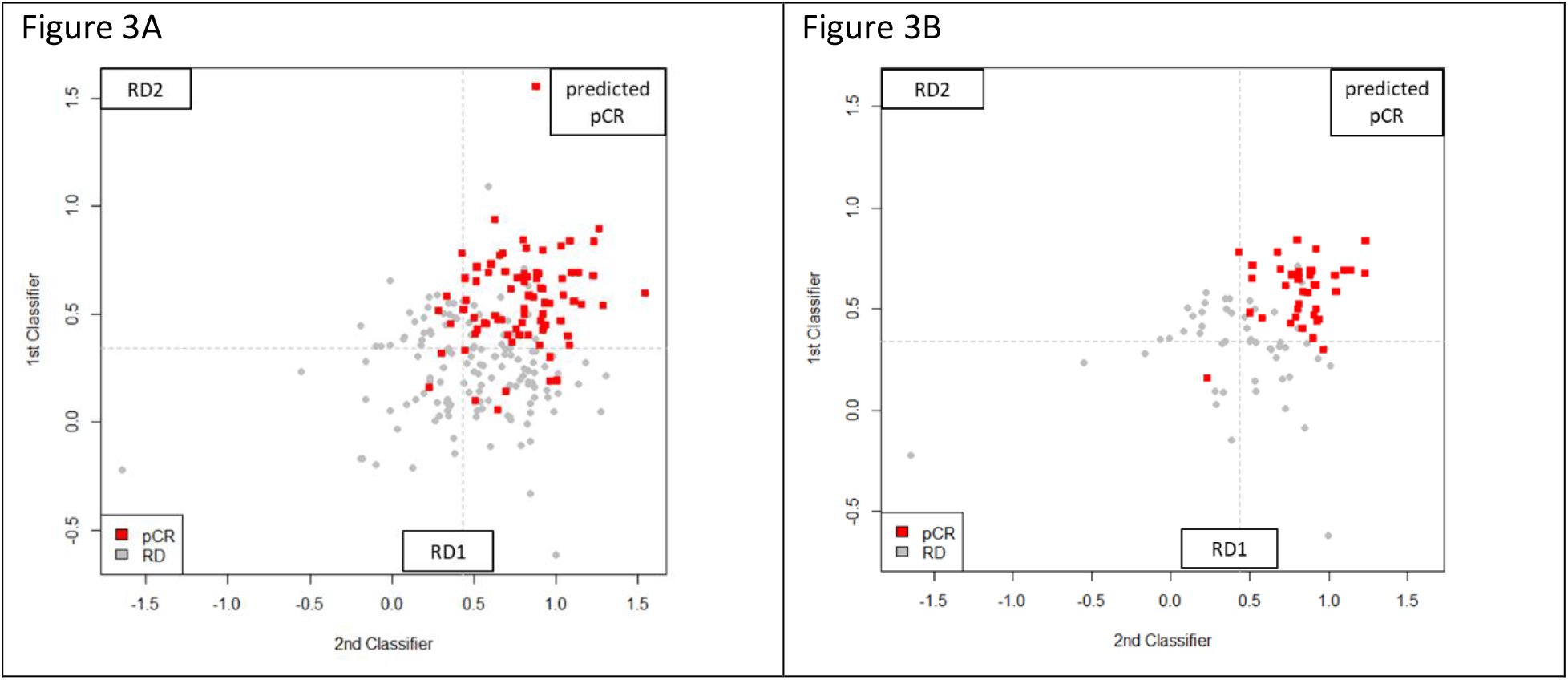
Output of the prediction scores for the total population of total 222 patients (A) and the 90 TNBC patients (B). The red squares represent patients achieving pCR, the grey squares are those with RD, and the dashed lines refer to the cutoff values above which pCR is predicted. Scores from Classifier 1 (18-gene) are on Y axis and Classifier 2 (15-gene) on the X axis. Patients predicted pCR by both classifiers are in the upper right quadrant. Patients predicted RD by the Classifier 1 are in the bottom half and patients predicted RD by Classifier 2 are in upper left quadrant. RD1 and RD2 represent patients with different biology based on the gene classifier used to stratify them.

**Table 2** shows the results of applying the 18-gene model and 15-gene model to stratify our 222 patients into pCR, and RD (RD1 and RD2 combined). The pCR rates of each population are shown to range from 18.84% for ER+/HER2-to 46.03% in HER2+ patients. The test correctly stratified 83.75% (67/80) of pCR and 86.62% (123/142) of RD patients in the total population, and 92.10% (35/38) of pCR and 80.77% (42/52) of RD patients in the TNBC subset. The overall accuracy was 85.58% in the total cohort and 85.56% in the TNBC subset.

**Table 2.** Test performance metrics

| **222 Member Data Set** |  |  |  |  |  |  |  |  |  |  |
| --- | --- | --- | --- | --- | --- | --- | --- | --- | --- | --- |
|  | Actual population statistics | Stratification by 2 classifiers performance metrics |  |  |  |  |  |  |  |  |
|  | pCR | PPV | NPV | Sensitivity | Specificity | TP | FP | TN | FN | Total |
| ER+HER2- | 18.84% | 58.8% | 94.2% | 76.9% | 87.5% | 10 | 7 | 49 | 3 | 69 |
| TNBC | 42.22% | 77.8% | 93.3% | 92.1% | 80.8% | 35 | 10 | 42 | 3 | 90 |
| HER2+ | 46.03% | 91.7% | 82.1% | 75.9% | 94.1% | 22 | 2 | 32 | 7 | 63 |
| Total Population | 36.04% | 77.9% | 90.4% | 83.8% | 86.6% | 67 | 19 | 123 | 13 | 222 |

PPV: positive predictive values
NPV: negative predictive values
TP: true positives
FP: false positives
TN: true negatives
FN: false negatives

### Gene expression, and Clinical Data Analysis Among the Two RD Subsets

We applied Kaplan-Meier (KM) curves to investigate the differences in recurrence-free survival (RFS) among the patients who have achieved pCR or RD after NAC (Figure 4A), and with predicted pCR, RD1 and RD2 classification (Figure 4B). Concordant with prior reports, patients achieving pCR have better survival chances than patients with RD (N_pCR_ = 42, N_RD_ = 67, hazard ratio = 3.8, p = 0.06). RD2 patients show a significant worse prognosis in comparison with RD1 patients (N_pCR_ = 43; N_RD1_ = 46; N_RD2_ = 20, hazard ratio = 3.035, p = 0.098) (Figure 4B) suggesting that the rationale application of the 2 classifiers stratified RD patients into two groups with different biology and risk profiles.

**Figure 4:**
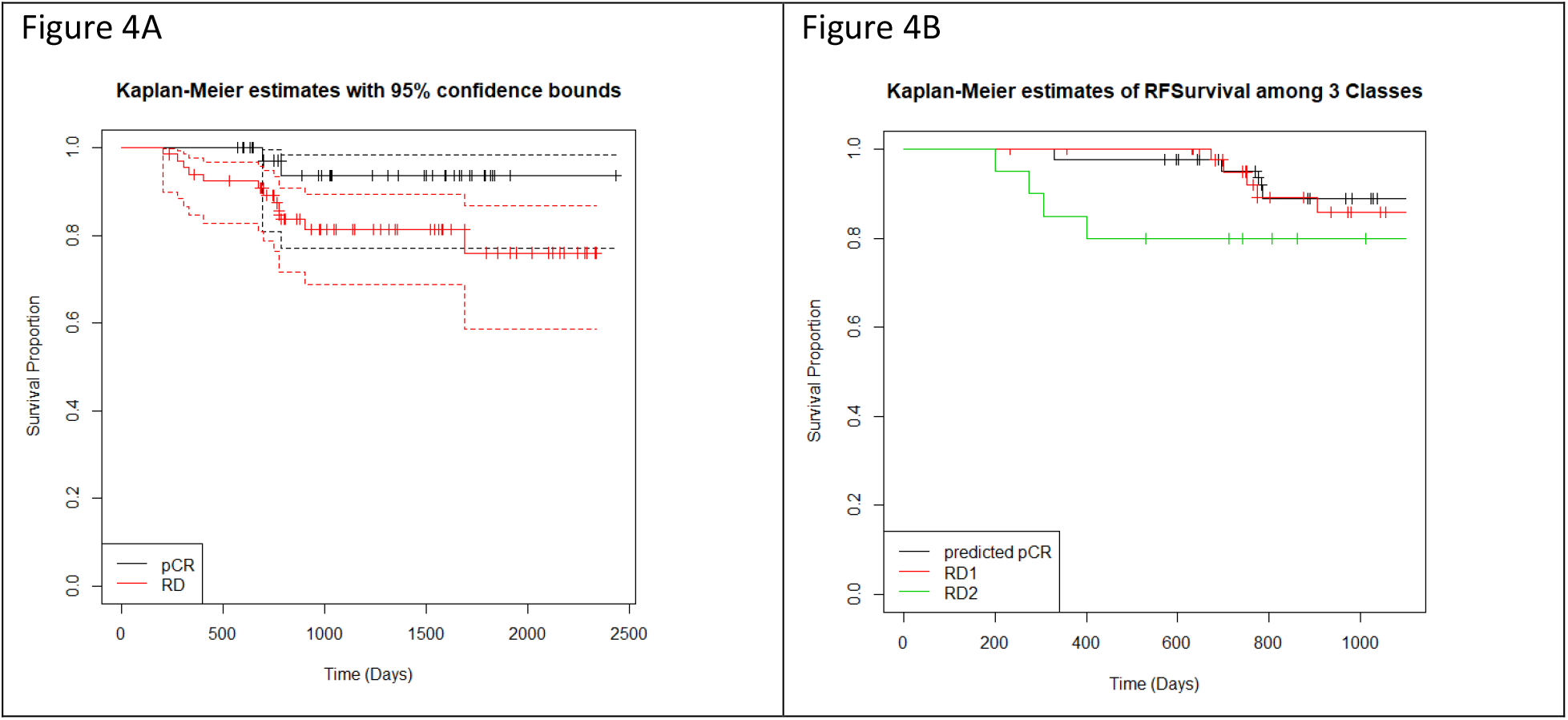
Kaplan-Meier curves showing RFS for 109 patients over a maximum of 4 years of follow up. (A) RFS separated by actual pCR (black line) and RD (red line) prior to stratification by the two classifiers in a subset of 109 patients (pCR=42, RD=67) for which data was available. The 95% confidence intervals are indicated by the dashed lines. (B) RFS for each of the three Classes following stratification by the two classifiers (pCR, RD1, and RD2).

Next, we investigated the expression of the proliferation marker Ki-67 and Androgen Receptor (AR) in triple negative breast cancer samples predicted pCR or RD (RD1 and RD2) (Figure 5). Figure 5A shows that while box plots did not show statistical difference in Ki-67 expression between TNBC predicted pCR and RD2, each with median expression equals to 5.2 and 4.6, respectively, RD1 samples are found to have significantly lower levels than the other classes with a median expression of 4.13 and an interquartile range of 2.79 to 4.76 (pCR vs ClRD1 p= 5.019e-05). We have also assessed AR expression, and as shown in Figure 5B, results RD1 had a statistically significant higher median expression of AR, with a median expression of 1.08 and an interquartile range of −0.444 to 2.433 in comparison to RD2 (RD1 vs RD2 p= 0.008403). Hence RD1 subgroup has the lowest KI67 and highest AR expression.

**Figure 5:**
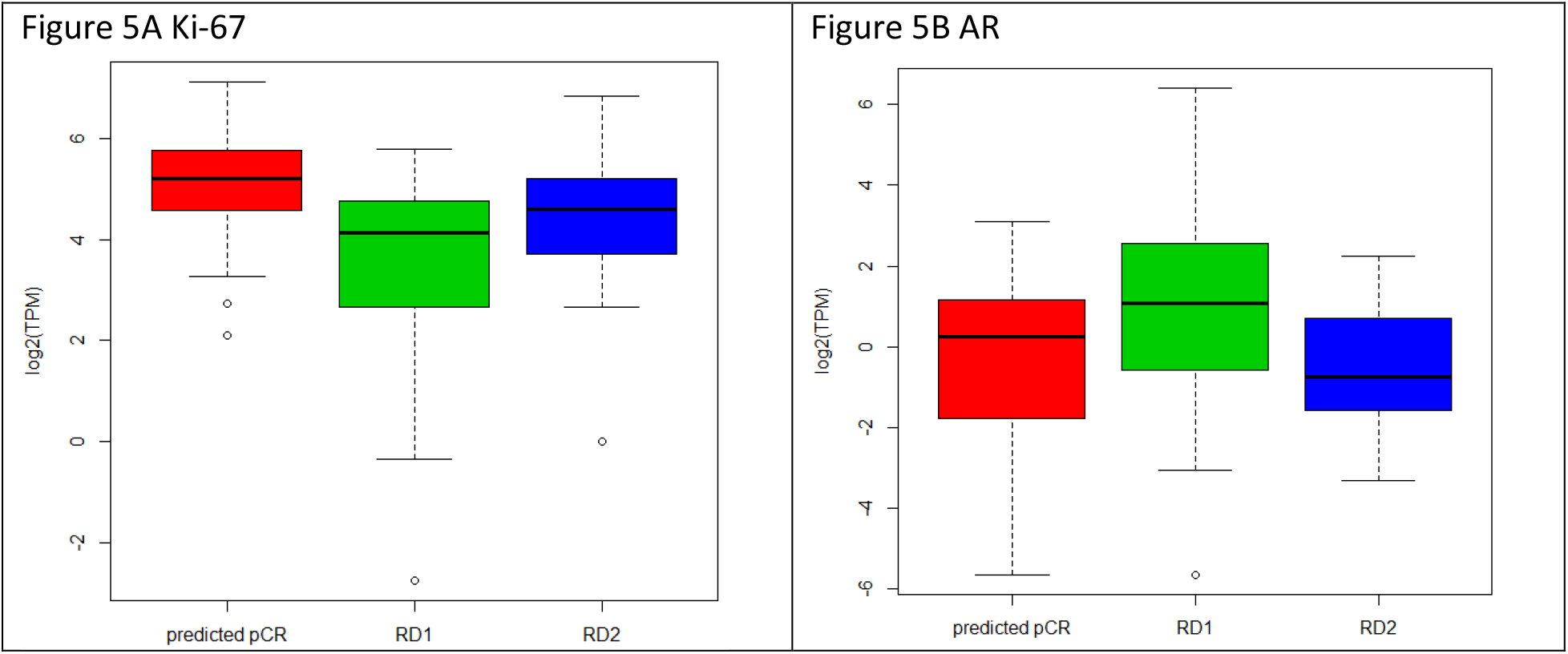
Gene expression and tumor grade comparisons with TNBC classes. A. The expression levels of Ki-67 are shown as a box plot for pCR (red), RD1 (green), and RD2 (blue), with each box representing the interquartile range of gene expression, and a horizontal line inside showing the median expression. Y-axis shows the Log2-transformed TPM values from RNA-seq expression. B. Expression of androgen receptor in TNBC tumors for each class is shown as in A.

Since RD2patients had the worst survival rate, we tested the correlation between our classifications and tumor stages and tumor grades obtained from the pathology reports (Figure 6). It was observed that TNBC patients predicted pCR contained over 60% stage 2 patients and about 30% stage 3 patients. TNBC patients predicted RD (RD1nad RD2) contained a decreased number of stage 2 (50%) and increased stage 3 (40%) cases respectively compared to the pCR group. There were more stage IV patients in RD2 in comparison with RD1(1.05% in RD1, and 6.67% in RD2) (Figure 6A). In regard to tumor grade, as expected from the Ki-67 and AR data, pCR patients contained the highest percentage of grade 3 patients (83.5% in pCR, 38.7% in RD1, and 63.3% in RD2), confirming that higher grade predicts for enhanced response to NAC (Figure 6B). The majority of the grade 1 patients (14 out 16) were found in RD1, while RD2 contained no grade 1 patients. Grade distribution of RD2 patients was intermediate between the pCR and RD groups but had worse prognosis, confirming the difficulty in predicting outcomes by clinical/pathologic data like grade or mitotic index.

**Figure 6:**
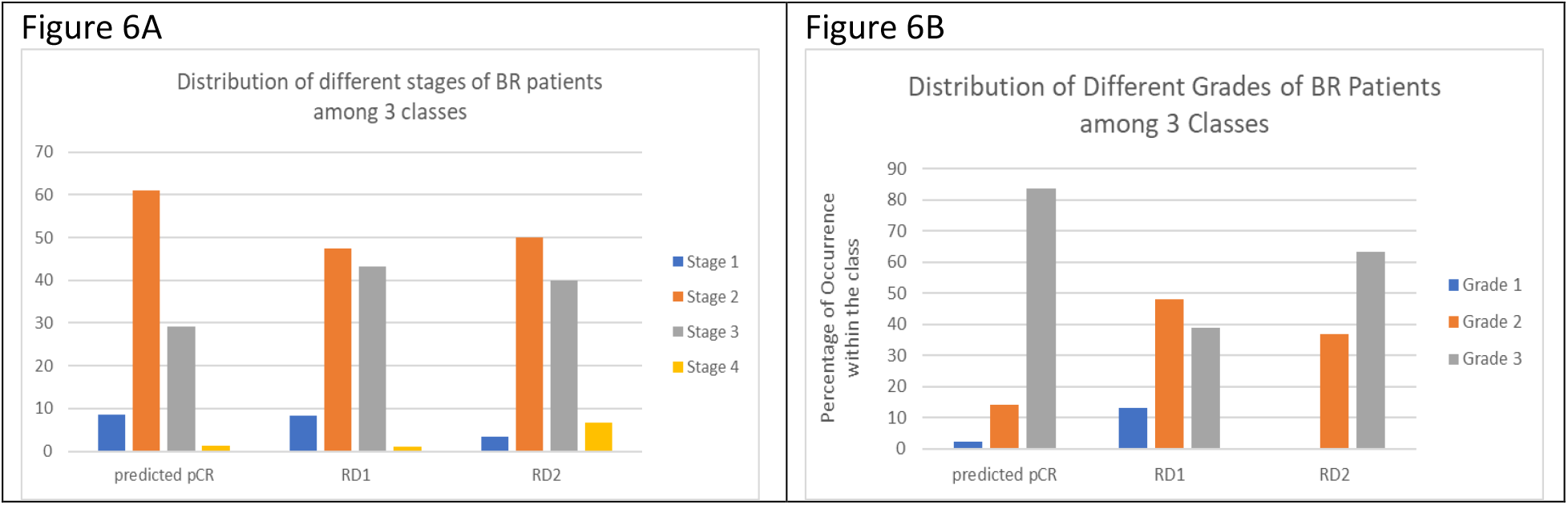
Tumor stage and tumor grade comparisons in TNBC classes. A. Bar plots showing the distribution of different stages of TNBC patients among the 3 classes (pCR, RD1, and RD2). Y-axis shows the percentages of patients at a given stage of breast cancer within each class. B. Bar plots showing the distribution of different grades of TNBC patients among the 3 classes as described in A.

## Discussion

We described here the development of a predictive test for pCR to NAC based on mRNA expression of genes analyzed using the clinically accessible technology of AmpliSeq. With the TNBC population, it is well-known that achieving a pCR is an accepted surrogate marker for survival, while RD is indicative of a likelihood for patient poor outcome [[2], [6]]. Since some patients will achieve pCR following standard therapy while others will require more intensive regimens, the ability to predict pCR, and as importantly RD, will impact physician choice and personalize treatment approaches for the individual patient. Incorporation of biomarker-based classifiers for outcome prediction is needed to improve overall patient outcomes by allowing tailoring of treatments in advance and improving outcomes with a reduction in the therapeutic toxicity and financial burden incurred by patients [[26], [27]]. TNBC patients with RD after NAC are six times more likely to have recurrence and twelve times more likely to die of metastatic disease [7]. Thus, accurate prediction of patients who are likely to have RD before standard NAC will allow the selection of therapeutic approaches based on patient biology.

We have previously reported RNA expression profiling using the publicly available microarray data sets collected from initial biopsy samples of breast cancer patients, to predict pCR or RD in response to NAC [14]. Due to the rapidly increasing application of next generation sequencing (NGS) technology for cancer diagnostic testing, we have used AmpliSeq to profile the biopsy samples of a validation unpublished cohort of stage I-IV breast cancer patient FFPE samples. The AmpliSeq data were then analyzed using a rationale sequential application of RNA classifiers to predict pCR and RD with 90% negative predictive and 78% positive predictive value in the overall cohort, and an even higher negative predictive value of 93%, and 78% positive predictive value for the TNBC patients. In our previous work, we have shown that achieving pCR was associated with significantly improved DRFS in TNBC patients [14]. In the current work, we have further demonstrated that patients achieving pCR displayed significantly better relapse-free survival compared to the predicted RD patients (Figure 4) which is consistent with our prior work and extensive literature.

Our classifiers have further stratified patients with residual disease into 2 classes (RD1 and RD2) with distinct biological features and significant differences in RFS (Figures 4). We have observed that RD1(lower risk RD) exhibits significantly lower expression of Ki-67 and higher androgen receptor expression than the higher-risk RD2 or predicted pCR classes. The breakdown of RD patients into a high and lower risk group of relapse and death is important not only in scientific investigation of the drivers of lack of response and progression but also the management of these patients in terms of therapeutic choices.

As shown in our prior study, [14] the rationale sequential application of 2 classifiers allowed the discovery of a subset of patients predicted to have RD with distinct biology and clinical profiles. While the 2 RD groups showed differences in biology and outcome they could not be distinguished based on any single factor such as tumor stage, tumor grade or Ki-67 and AR expression levels indicating the importance of incorporation of additional parameters to evaluate patient prognosis and response to treatments.

Lehmann et al (2016) [18] has previously shown that TNBC is a highly diverse type of breast cancers and defined four TNBC subtypes based on a retrospective analysis of gene expression datasets from five clinical trials [5]. In combination with TCGA genomic and clinical data, the authors showed that TNBC subtypes significantly differ in response to similar NAC achieving pCR in 41% of the Basal-like BL1-subtype (low AR but elevated cell cycle and DNA damage response) compared to 18% for Basal-likeBL2-subtype and 29% for Luminal Androgen Receptor LAR-subtype (highest expression of AR). A new three-subtype classification of TNBC has recently been published in *Breast Cancer Research* [28]. In their study identification of 3 molecular clusters; C1 with apocrine features, luminal and PIK3A-mutated. C2 and C3 with basal-like features. These classes show remarkable similarities in terms of androgen receptor and Ki-67 expression/cell cycle processes, to our classifier. Their classifier looked at links between neurogenesis, tertiary lymphoid structures, plasma cells, B lymphocytes, and triple negative breast cancer subtypes. Ours on the other hand was developed utilizing markers of non-malignant breast epithelial cell organization. How our biomarker array and their classification might overlap in terms of gene expression and response to NAC is unclear.

Our RNA-profiling strategy in pretreatment TNBC allows identification of TNBC patients that will likely respond and to those who will not respond to standard of care NAC utilizing anthracyclines and taxanes. It therefore allows reasonable utilization of more experimental and or toxic treatment regimens in patients predicted to not respond. The identification of RD2 high risk patients will have significant impact on the TNBC diagnosis and treatment. However, the genetic/immunologicfactors underlining the worst survival rates of the RD2 patients are still not clear. Obtaining the genomic data including mutation, copy number loss or amplification, as well as gene fusion for the 222 patients might help explain why RD2 patients have the worst survival compared to the RD1 and pCR patients. Additionally, driving down to immune response markers utilized in the Jezequel study [28] would be of great interest and may define the place for immunotherapy in these patients.

## Materials and Methods

### Patients and Samples

Two hundred and eighty-seven breast cancer patient samples were obtained from multiple clinical sites via the MT Group, UConn Health, Hartford Hospital and the FIOCRUZ institute (Brazil). All samples were obtained under IRB review and were waived from consent. UConn Health, Hartford Hospital and FIOCRUZ institute sample collection was approved under UConn IRB #17-013-6.1. Patient inclusion criteria: female patients between 21 to 90 years of age; patients with a biopsy-proven breast cancer diagnosis; patients received chemotherapy in a neoadjuvant setting; patient samples contained sufficient breast cancer cells in pathology tissue block; patients were diagnosed with breast cancer between the year 2004 and the present. Patient exclusion criteria: patient samples lacking sufficient tumor tissue (<30%) upon pathology review. Sample specifications: Breast tumor biopsy and lymph node biopsy (when available) collected prior to patient receiving neoadjuvant taxane-based chemotherapy per their physician’s discretion and provided as formalin fixed, paraffin embedded (FFPE) tissues. Upon pathological examination, the samples belonged to Stages I-IV and T1-T4, N1-N3, M0-M1 breast cancer. Relevant pathologic, clinical, and demographic parameters were extracted from de-identified patient pathology reports for use in model development. Pathologic Complete Response and Residual Disease were defined by the block’s Institution of origin in the pathology report, and the annotation was used for the study.

Relevant pathologic, clinical, and demographic parameters were extracted from de-identified patient pathology reports for use in model development.

### RNA and library preparation for NGS

Total RNA was extracted from FFPE samples and quantified using Qubit4 system. AmpliSeq library preparation was performed according to the AmpliSeq for Illumina Transcriptome Human Gene Expression Panel workflow. Library quality control analysis was performed on the Agilent TapeStation System. Sample libraries passing TapeStation QC were quantified using the Qubit 4 and then normalized, pooled, and loaded into the Illumina NextSeq 500/550 High output 300 cycle v2.5 kit for sequencing of 2×150 paired-end AmpliSeq.

### Data set for predictive model development

A total of 291 clinical samples, from 279 unique patients were sequenced using AmpliSeq of the Illumina NGS platform. 4 patients were sequenced twice, which led to 287 unique samples in total. Among the 287 sample which have been sequenced, 23 of them did not pass the QC criteria (based on total number of output reads obtained from each sample, percentage of mapping rates, as well as percentage of pairing from paired-end reads), thus were excluded from subsequent analysis, leading to 264 samples, which could be used for further analysis. These 264 samples corresponded to 253 unique patients, since there were 11 patients who contained two biopsy samples. The two biopsies were taken either from two separate breast tissue sites, or one from breast tissue and one from lymph nodes, resulting in 22 unique clinical samples. Two hundred and twenty two out of 264 sequenced samples had the clinical data available, which were obtained from various clinical sites or hospitals, including the MT group, UConn Health, Hartford Hospital, and FIOCRUZ Institute (Brazil). The RNA expression data from these 222 samples was used to further analyses.

### AmpliSeq Data Processing

Raw fastq files from sequencing runs were processed as follows: Atropos was used to remove the adapter sequences from all the reads; STAR aligner was used to map the reads to the human hg38 genome; RSEM was used to quantitate the reads for each individual gene. Finally, the TPM values were used to represent the mRNA expression of each individual genes. In our first AmpliSeq experiment, we did an experimental study on breast cancer samples at 3 dilutions, and analyzed the results using different aligners, gene quantitation methods, and read count normalizations. The combination that gave the most reproducible results was STAR/RSEM/TPM.

### Development of RNA-biomarker classifiers

RNA classifiers of 18-genes and 15-genes were developed following methods previously described [14]. Briefly, the initial 222-patient cohort was divided into a training and a testing set with the 80%/20% regimen, and each set contains similar fractions of pCR and RD patients, as well as having the similar representations of ER, HER2 and PGR statuses. The BRGLM algorithm was used to select the genes having the most pCR predictive power, which resulted in the 18-gene and 15-gene classifiers. The first classifier (18-gene) stratified the patient cohort into predicted pCR and predicted RD groups respectively. The second classifier (15-gene) was sequentially applied to the predicted pCR patients only and further stratified a second group of predicted of RD patients (Figure 1). Therefore the two classifiers applied in rational sequence stratified two biologically distinct classes of RD patients (RD1 and RD2) while decreasing false positive rates. The genes comprising the classifiers are shown in Supplemental Table 1 along with their coefficients, intercepts, and threshold values above which pCR is predicted.

### Kaplan–Meier Survival Analysis

The survival data were collected for 109 patients that had information for first visit, last visit, and date of relapse annotated. The relapse-free survival (RFS) was calculated as the total number of days between the first visit and last visit for patients that did not relapse, or as the total number of days between the first visit and the date when relapse occurred, for patients with disease recurrence.

## Data Availability

The data will become available at GEO on March 1, 2021

