## Supplemental data for "RNA expression classifiers from a model of breast epithelial cell organization to predict pathological complete response in triple negative breast cancer"

Suppl. Figure 1: Heatmap view of the cluster analysis of 348 RNA expression biomarkers across 222 breast cancer FFPE biopsy samples. Log2-transformed TPM values were used in the cluster analysis. Red, higher expression; blue, lower expression. Rows represent genes and columns represent samples.

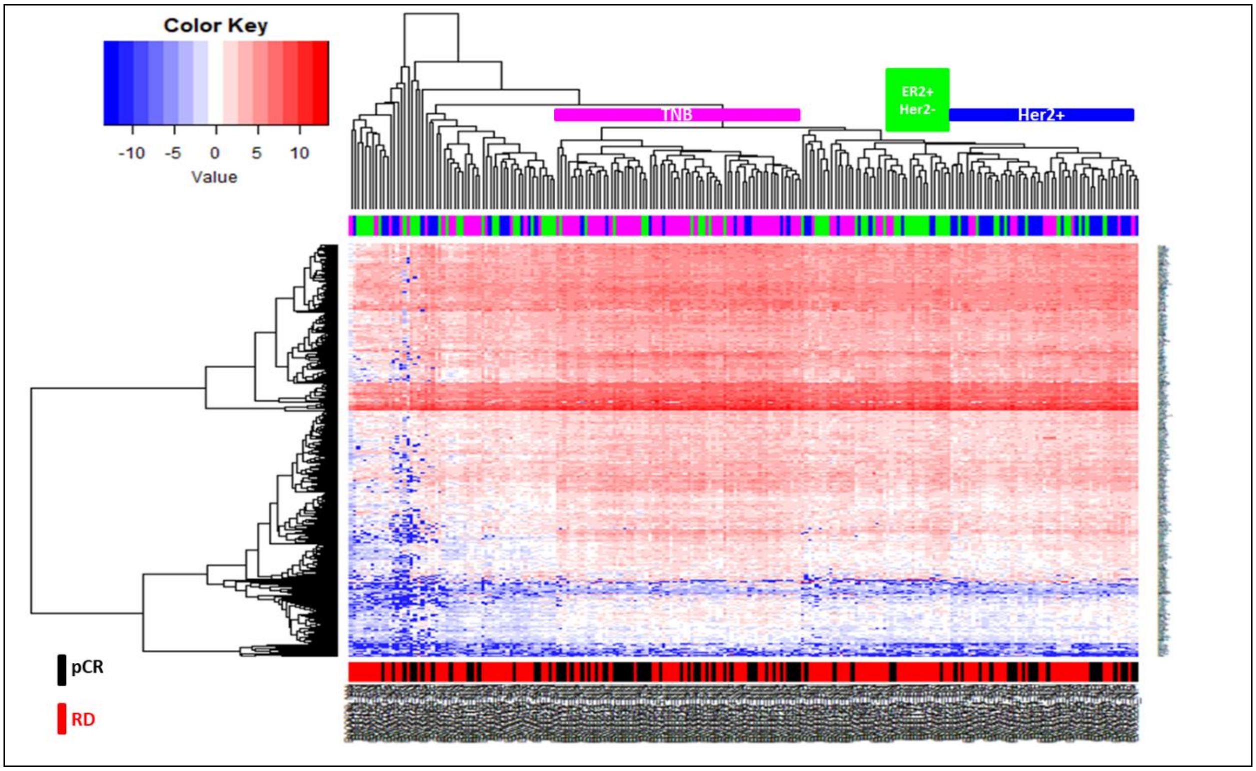

Suppl. Figure 2: Results of the BRGLM runs. A) Bar plot shows the occurrence of the 348 genes following 10,000 BRGLM runs with bootstrapped of samples and genes. Each bar represents the occurrence from one individual gene. B) Dot plot of the top 138 genes with the highest occurrence, sorted from the highest to the lowest.

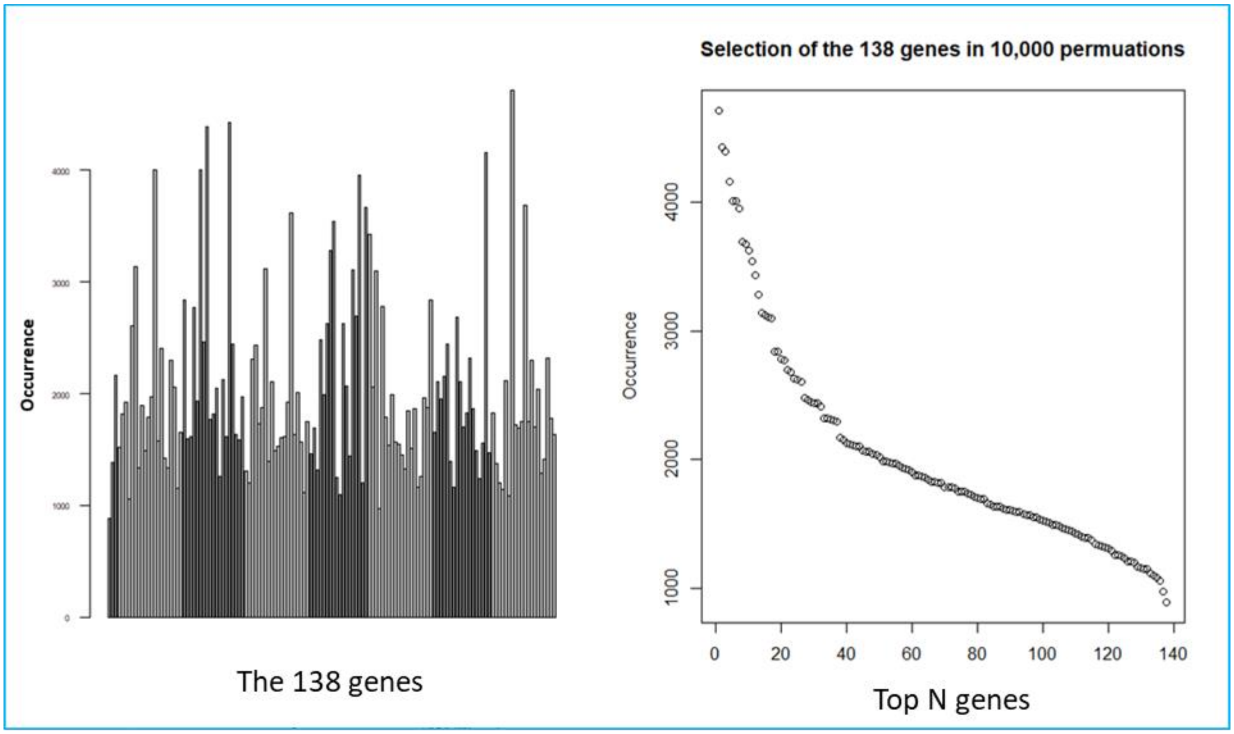

Suppl Table 1: The gene symbols, coefficients, intercepts, and threshold values are shown for Classifiers 1 and 2. Positive coefficients predict pCR and negative coefficients RD.

a) Classifier 1 (cutoff = 0.34328915)

| 18-gene model | 18gene$coef |
|---|---|
| (Intercept) | 0.60068646 |
| TPRKB | 0.04122403 |
| FEN1 | 0.09650187 |
| ELN | -0.05829091 |
| STK17A | 0.07627754 |
| DVL1 | -0.07539091 |
| CENPN | -0.02195906 |
| NOC2L | 0.11499943 |
| TTK | 0.04609686 |
| NUP153 | 0.12210722 |
| LRP8 | 0.03412527 |
| MRPL35 | -0.08945316 |
| NUP205 | 0.0613218 |
| MFAP4 | 0.05556132 |
| CAPRIN2 | -0.10383583 |
| HPCAL1 | -0.01683015 |
| NDC1 | -0.0079133 |
| NUSAP1 | -0.05668388 |
| RCC1 | -0.08360381 |

b) Classifier 2 (cutoff = 0.43332966)

| 15-gene model | 15gene$coef |
|---|---|
| (Intercept) | 0.761853 |
| EHF | -0.08809 |
| PGK1 | -0.18156 |
| DUSP6 | 0.129921 |
| BCAR3 | -0.12955 |
| CYP51A1 | -0.14268 |
| TPRKB | 0.008704 |
| SCD | 0.126011 |
| CENPN | -0.00293 |
| FEN1 | 0.063584 |
| NIF3L1 | 0.031227 |
| ALG8 | -0.0189 |
| IMPDH1 | -0.09656 |
| RANBP1 | 0.231282 |
| PRDX3 | -0.14077 |
| HAT1 | 0.114986 |
